# Greater intergroup bias in vaccination attitudes among physicians than the general public

**DOI:** 10.64898/2026.04.23.26351641

**Authors:** Michio Murakami, Fumio Ohtake

## Abstract

While vaccination conflicts have become apparent, physicians’ attitudes toward those with differing views remain unclear. Through an online survey of 492 physicians and 5,252 members of the general public in Japan in February 2026, we investigated attitudes toward four vaccines (influenza, measles, HPV, and COVID-19). Intergroup bias was assessed as ingroup minus outgroup attitudes using a feeling thermometer. Multilevel regression examined associations with agreement group and physician status. Intergroup bias was significantly positive in both agreement and disagreement groups across all vaccine types, and was higher in the agreement group. Physicians exhibited higher intergroup bias than the general public. These findings indicate that vaccination conflict is bidirectional: physicians, often viewed as targets of hostility from vaccine-hesitant individuals, themselves exhibit greater intergroup bias toward those with opposing views. Interventions to raise physicians’ awareness of their own bias, alongside communication strategies for vaccine-hesitant individuals, are needed.

## 1. Introduction

The coronavirus disease 2019 (COVID-19) pandemic has led to conflicts and social polarization regarding vaccination attitudes [1]. While promoting effective vaccines is crucial for preventing infectious diseases, some individuals hesitate to vaccinate for various reasons, including medical concerns and religious beliefs. COVID-19 vaccine opponents have been reported to adopt strongly oppositional stances and engage in hostile behavior toward supporters on social media [2]. Healthcare workers have also faced harassment and threats from vaccine opponents [3]. Conversely, in countries where COVID-19 vaccination has been mandated for healthcare workers, those opposing the mandate have experienced reputation damage or dismissal [4].

Previous studies have documented the knowledge gaps and conflicts between vaccine supporters and opponents. Individuals who strongly oppose vaccines tend to have low objective knowledge yet perceive themselves as knowledgeable [5]. On social media, although opponents are fewer in number, they dominate their networks, whereas supporters are less likely to engage in discussions [6]. Vaccine supporters report stronger negative or discriminatory attitudes toward the unvaccinated or opponents than vice versa [7, 8]. However, no studies have examined discriminatory attitudes among healthcare professionals toward groups with differing vaccination views, or differences between physicians and the general public in such conflict. Given that physicians strongly influence vaccination decisions [9, 10], it is important to clarify the physicians’ attitude toward group with differing views, as their recommendation can be biased by patient characteristics, including their socioeconomic status and their vaccination stance [11].

Therefore, this study investigated attitudes toward vaccination for four infectious diseases, including seasonal influenza, measles, human papillomavirus (HPV), and COVID-19, which differ in vaccine efficacy, as well as attitudes toward groups holding opposing views, among physicians and the general public.

## 2. Methods

### 2.1. Ethics

This study was approved by the Ethics Committee of the Center for Infectious Disease Education and Research at the University of Osaka (approval numbers: 2025CRER0120; 2026CRER0415-2). Informed consent was obtained from all participants. Participants received reward points redeemable for merchandise.

### 2.2. Participants

We conducted online surveys of physicians and the general public aged 20–69 in Japan from February 17–18, 2026, and from February 17–25, 2026, respectively. All participants were registered monitors with Nikkei Research Inc. Physicians’ status was verified through an authentication system including university graduation records and workplace mail verification; approximately two-thirds of physicians in Japan are registered.

Target sample sizes were 440 physicians and 5,000 individuals from the general public (8 regions × 625 respondents). Although the required sample size was 384 based on a 95% confidence interval (CI) and a 5% margin of error [12], larger sample sizes were set to ensure sufficient power. Data from the Japanese general public were collected by region to match the national age and gender distribution.

Of the 1,041 physician respondents, 549 were excluded (18 without consent, 66 outside the age range, 13 with unspecified gender, 24 dropouts, and 428 due to quota completion), leaving 492. Among 6,794 general public respondents, 1542 were excluded (405 without consent, 4 outside the age range, 26 with unspecified gender, 1 outside Japan, 793 dropouts, and 313 due to quota completion), resulting in 5,252 participants.

### 2.3. Vaccines

We analyzed four vaccines: seasonal influenza, measles, HPV, and COVID-19. The following summary of efficacy and current recommendations draws on the corresponding World Health Organization documents [13-16]. Influenza vaccine efficacy varies by strain match (e.g., 59% efficacy against clinical illness over 12 seasons among adults aged 18–65) [13]. Two doses of measles vaccine provide high lifelong immunity (approximately 95%) [14]. HPV vaccination provides long-term protection (over 10 years) with high efficacy (e.g., 88% reduction in cervical adenocarcinoma in females aged 15–26) [15]. For COVID-19, booster doses are recommended for older adults and those with underlying conditions due to waning protection against infection but sustained protection against severe disease, particularly after the emergence of the Omicron variant [16].

In Japan, all vaccinations are voluntary. As of the February 2026 survey, Influenza vaccines are administered annually, measles vaccines in two routine childhood doses, HPV vaccines in two to three doses for girls aged 12–16, and COVID-19 vaccines annually for adults aged ≥65 years and high-risk individuals aged 60–64.

### 2.4. Survey items

#### 2.4.1. Outcome

Intergroup bias was used as an indicator of antagonistic feelings between groups. It was calculated from participants’ agreement with vaccination and their attitudes toward those who agree or disagree [8]. Participants rated agreement with each vaccine on an 11-point Likert scale (0 = strongly disagree to 10 = strongly agree). Attitudes toward others were assessed using a feeling thermometer (0 = strongly dislike to 10 = strongly like). Scores of 6–10 defined the agreement group and 0–4 the disagreement group (excluding 5 as neutral). Intergroup bias was calculated as ingroup minus outgroup attitudes. The ingroup comprised those sharing the participant’s stance, and the outgroup the opposite. Higher values indicate stronger ingroup preference.

#### 2.4.2. Independent variables

Independent variables were vaccination agreement group and participant type (physician vs. general public). Physicians also reported specialty (internal medicine, pediatrics, psychiatry, surgery, or other).

Covariates included perceived benefits (two items on infection and severe disease prevention), perceived risks (two items on mild and severe adverse events), age, gender, presence of children, and partner status. All perception items used 11-point Likert scales. These variables were included based on prior associations with vaccination behavior [17, 18] and to assess whether differences in intergroup bias persisted after adjustment. The survey also collected additional variables (e.g., trust in information sources, health literacy, recommendation intentions, infection control perceptions, vaccination history, COVID-19 history, and sociodemographic factors), which are beyond the scope of this study.

Two directed question scale (DQS) items were used to identify inattentive respondents [19], with warnings provided upon failure. Respondents failing either DQS item, general public respondents reporting “physician” as occupation, and physicians aged <24 years were classified as inattentive or ineligible. Participant characteristics are shown in Table 1.

**Table 1.**
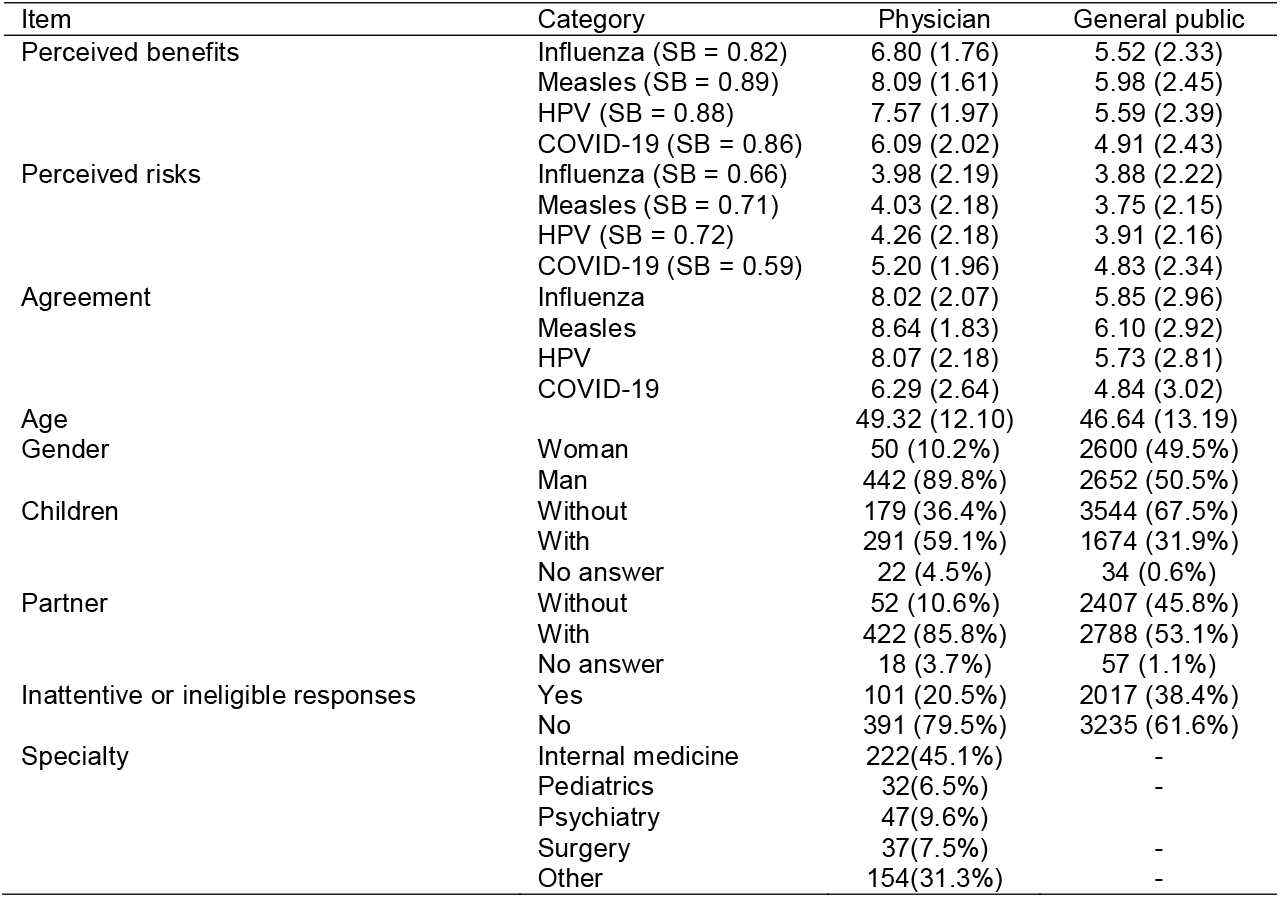
Participant characteristics. Values are mean (standard deviation) or n (%). SB: Spearman–Brown coefficient.

### 2.5. Statistical analysis

For each vaccine, 95% CIs for intergroup bias, ingroup attitudes, and outgroup attitudes were calculated separately by agreement group and participant type (physicians vs. the general public). In main analysis 1, multilevel regression models were fitted with intergroup bias, ingroup attitudes, or outgroup attitudes as outcomes, and agreement group and participant type as independent variables. Covariates included perceived benefits, perceived risks, agreement magnitude, age, gender, children, partner status, and vaccine dummy variables. Average scores were used for perceived benefits and risks after confirming high internal consistency. For perceived benefits, risks, and agreement magnitude, distance from the neutral midpoint (5) was used. Participant ID was included as a random effect.

In main analysis 2, analyses were restricted to physicians, with agreement group and specialty as independent variables.

Sensitivity analyses for main analysis 1 included: (1) standard multiple regression stratified by vaccine type; (2) multilevel models stratified by agreement group using absolute values instead of distance from neutral; and (3) analyses excluding inattentive or ineligible respondents.

Variance inflation factors confirmed no substantial multicollinearity. Analyses were performed using IBM SPSS version 28.

## 3. Results

For seasonal influenza, measles, HPV, and COVID-19, the proportions of physicians in the agreement group were 88.2%, 92.9%, 87.4%, and 63.8%, respectively, compared with 51.8%, 54.3%, 48.1%, and 38.6% in the general public. Intergroup bias was significantly positive in both agreement and disagreement groups. In the physician disagreement group, point estimates were also positive but did not reach statistical significance, likely reflecting the small sample size (Table 2; e.g., measles: physicians—disagreement 1.71 [95% CI −0.17 to 3.60], agreement 5.60 [5.27–5.92]; general public—disagreement 0.48 [0.31–0.64], agreement 3.21 [3.09–3.34]).

**Table 2.**
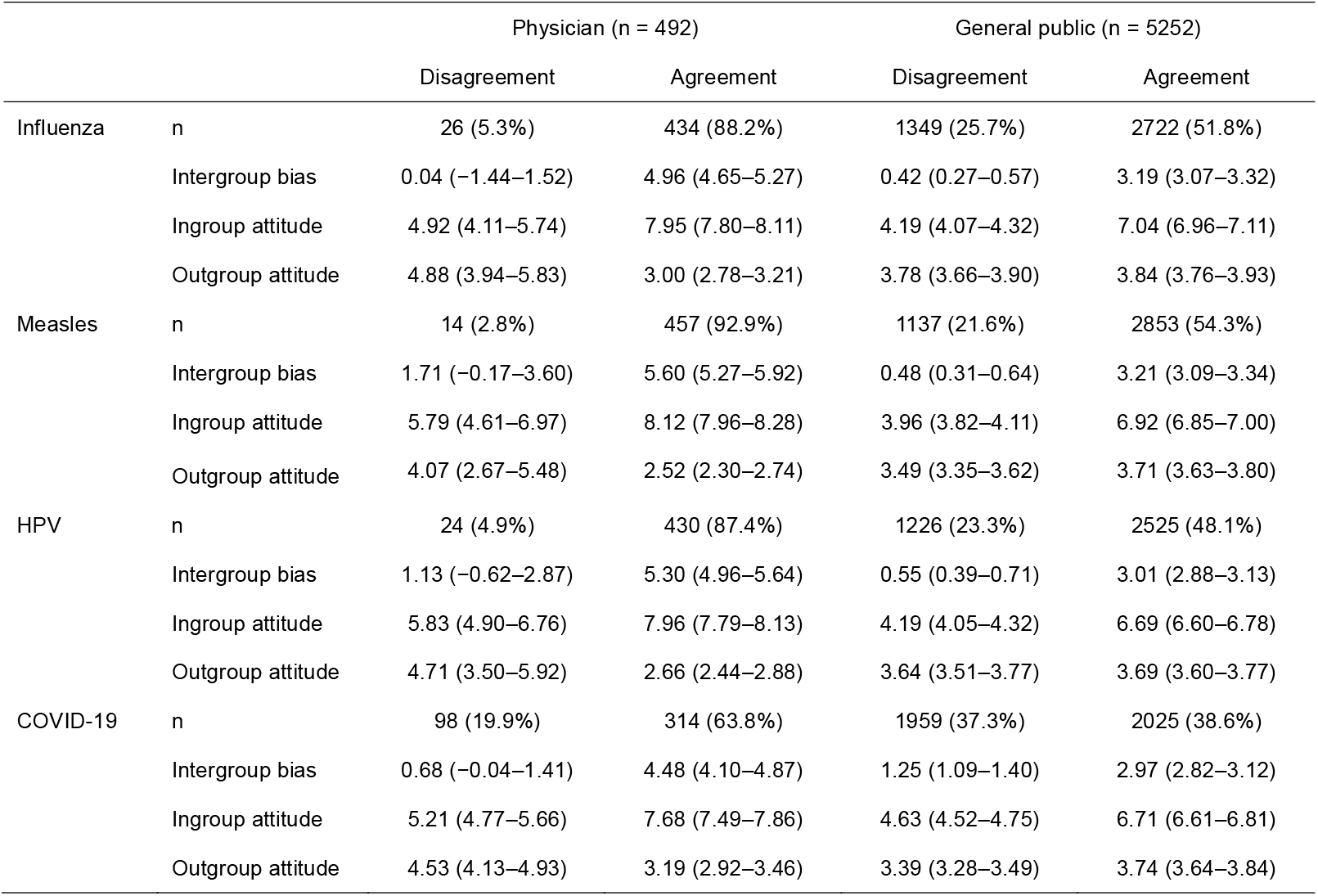
Intergroup bias, ingroup attitudes, and outgroup attitudes regarding vaccination agreement among physicians and the general public. Values are mean (95% confidence intervals) or n (%).

|  |  | Physician (n = 492) |  | General public (n = 5252) |  |
| --- | --- | --- | --- | --- | --- |
|  |  | Disagreement | Agreement | Disagreement | Agreement |
| Influenza | n | 26 (5.3%) | 434 (88.2%) | 1349 (25.7%) | 2722 (51.8%) |
|  | Intergroup bias | 0.04 (−1.44–1.52) | 4.96 (4.65–5.27) | 0.42 (0.27–0.57) | 3.19 (3.07–3.32) |
|  | Ingroup attitude | 4.92 (4.11–5.74) | 7.95 (7.80–8.11) | 4.19 (4.07–4.32) | 7.04 (6.96–7.11) |
|  | Outgroup attitude | 4.88 (3.94–5.83) | 3.00 (2.78–3.21) | 3.78 (3.66–3.90) | 3.84 (3.76–3.93) |
| Measles | n | 14 (2.8%) | 457 (92.9%) | 1137 (21.6%) | 2853 (54.3%) |
|  | Intergroup bias | 1.71 (−0.17–3.60) | 5.60 (5.27–5.92) | 0.48 (0.31–0.64) | 3.21 (3.09–3.34) |
|  | Ingroup attitude | 5.79 (4.61–6.97) | 8.12 (7.96–8.28) | 3.96 (3.82–4.11) | 6.92 (6.85–7.00) |
|  | Outgroup attitude | 4.07 (2.67–5.48) | 2.52 (2.30–2.74) | 3.49 (3.35–3.62) | 3.71 (3.63–3.80) |
| HPV | n | 24 (4.9%) | 430 (87.4%) | 1226 (23.3%) | 2525 (48.1%) |
|  | Intergroup bias | 1.13 (−0.62–2.87) | 5.30 (4.96–5.64) | 0.55 (0.39–0.71) | 3.01 (2.88–3.13) |
|  | Ingroup attitude | 5.83 (4.90–6.76) | 7.96 (7.79–8.13) | 4.19 (4.05–4.32) | 6.69 (6.60–6.78) |
|  | Outgroup attitude | 4.71 (3.50–5.92) | 2.66 (2.44–2.88) | 3.64 (3.51–3.77) | 3.69 (3.60–3.77) |
| COVID-19 | n | 98 (19.9%) | 314 (63.8%) | 1959 (37.3%) | 2025 (38.6%) |
|  | Intergroup bias | 0.68 (−0.04–1.41) | 4.48 (4.10–4.87) | 1.25 (1.09–1.40) | 2.97 (2.82–3.12) |
|  | Ingroup attitude | 5.21 (4.77–5.66) | 7.68 (7.49–7.86) | 4.63 (4.52–4.75) | 6.71 (6.61–6.81) |
|  | Outgroup attitude | 4.53 (4.13–4.93) | 3.19 (2.92–3.46) | 3.39 (3.28–3.49) | 3.74 (3.64–3.84) |

After adjustment for covariates (main analysis 1), the agreement group remained positively associated with intergroup bias (unstandardized partial regression coefficient B = 1.82 [1.72–1.91]; Tables 3 and S1). Physician status was also independently associated with higher intergroup bias (1.66 [1.40–1.93]). Both the agreement group and physicians showed higher ingroup attitudes and lower outgroup attitudes.

**Table 3.**
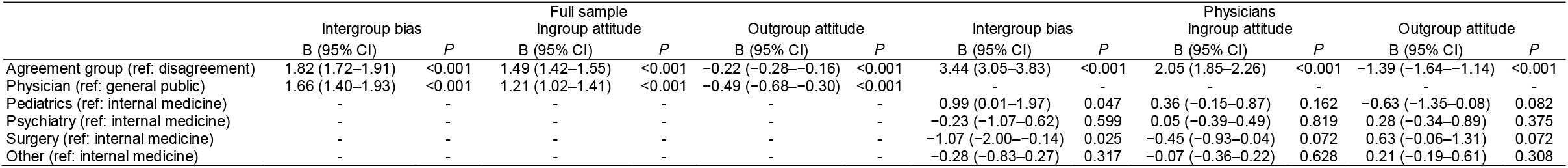
Unstandardized regression coefficients for intergroup bias, ingroup attitude, and outgroup attitude. Covariates included perceived benefits stance from neutral), perceived risks (distance from neutral), agreement (distance from neutral), age, gender, children, partner, and vaccine dummy riables. Full results are shown in Table S1. Random effects are as follows: full sample—intergroup bias, between-person standard deviation (σ_u_) =, residual standard deviation (σ_e_) = 1.74, intraclass correlation coefficient (ICC) = 0.66; ingroup attitude, σ_u_ = 1.85, σ_e_ = 1.06, ICC = 0.75; outgroup attitude, σ_u_ = 1.79, σ_e_ = 1.11, ICC = 0.72; physician—intergroup bias, σ_u_ = 2.48, σ_e_ = 1.68, ICC = 0.68; ingroup attitude, σ_u_ = 1.28, σ_e_ = 0.91, ICC = 0.67; outgroup attitude, σ_u_ = 1.83, σ_e_ = 1.08, ICC = 0.74. Variance inflation factor: ≤ 1.55 for full sample; ≤ 2.59 for physicians.

In main analysis 2 (physicians only), surgeons showed lower intergroup bias than internists, whereas pediatricians showed higher intergroup bias (Table 3; surgery: B = −1.07 [95% CI −2.00 to−0.14], *P* = 0.025; pediatrics: B = 0.99 [0.01–1.97], *P* = 0.047). As in main analysis 1, the agreement group was positively associated with intergroup bias and ingroup attitudes and negatively associated with outgroup attitudes.

Sensitivity analyses for main analysis 1 supported these findings. Results were consistent across vaccine types (Table S2), although intergroup bias for COVID-19 tended to be smaller. In sensitivity analysis 2, no difference between physicians and the general public was observed in the disagreement group, whereas a significant difference remained in the agreement group (Table S3). Results were unchanged after excluding inattentive or ineligible respondents (Table S4).

## 4. Discussion

In this study, we examined intergroup bias regarding four vaccines among physicians and the general public. Intergroup bias was positive in both agreement and disagreement groups across all vaccine types, and was higher in the agreement group. After adjustment for covariates, this pattern persisted, and intergroup bias was also higher among physicians. Both ingroup and outgroup attitudes were significantly associated with agreement group and physician status, indicating that intergroup bias reflects contributions from both components. These findings were consistent across sensitivity analyses.

Greater intergroup bias in the agreement group is consistent with previous studies in the general public [7, 8]. One explanation is that vaccination is viewed as a mutual aid “social contract,” in which individuals reward compliance and punish non-compliance, leading to negative perceptions of “free riders” [20]. A novel finding of this study is that physicians exhibited higher intergroup bias regardless of whether they agreed or disagreed with vaccination. Although physicians have been viewed primarily as targets of hostility from vaccine opponents [3], our findings suggest that vaccination conflict is bidirectional, with physicians themselves exhibiting substantial bias toward those with opposing views. This pattern suggests that intergroup bias may be stronger among physicians more directly involved in vaccination or patient consultation. When stratified by vaccine type, intergroup bias among physicians was lowest for COVID-19. Among the four vaccines, COVID-19 has relatively lower efficacy and a shorter duration of protection [13-16], which may be associated with less confrontational attitudes toward opposing groups. Physicians may exhibit more confrontational attitudes toward outgroups with differing views on vaccination for vaccines with higher efficacy.

These findings have several implications. First, intergroup bias was stronger in the agreement group than in the disagreement group, suggesting that the agreement group may play a substantial role in shaping conflict. The agreement group should recognize vaccination as a best-effort responsibility and avoid exacerbating conflict. Second, higher intergroup bias among physicians—particularly those involved in vaccination or patient consultation—may influence how they communicate recommendations. Implicit biases can affect clinical decision-making, including vaccine recommendations [11]. Physicians’ intergroup bias may weaken recommendations to those in the disagreement group. Addressing such bias may help improve communication with individuals who oppose vaccination. Addressing this bias through medical education and communication training may help mitigate the bidirectional nature of vaccination conflict.

This study has several limitations. First, participants were recruited from a survey panel, introducing potential selection bias; however, physician status was rigorously verified, and approximately two-thirds of physicians in Japan are registered. Second, the study was conducted in Japan, limiting generalizability to other cultural contexts. Third, intergroup bias was measured using a feeling thermometer [8], and does not capture actual behaviors or their severity.

Despite these limitations, this study provides the first evidence that physicians harbor greater intergroup bias in vaccination attitudes than the general public.

## Fundings

This study was supported by Japan Society for the Promotion of Science (JSPS) KAKENHI JP25H00388 and Topic-Setting Program to Advance Cutting-Edge Humanities and Social Sciences Research, and The Nippon Foundation–The University of Osaka Project for Infectious Disease Prevention.

### CRediT authorship statement

Conceptualization: Michio Murakami and Fumio Ohtake

Methodology: Michio Murakami and Fumio Ohtake

Formal analysis: Michio Murakami

Investigation: Michio Murakami and Fumio Ohtake

Data curation: Michio Murakami

Writing - Original Draft: Michio Murakami

Writing - Review & Editing: Fumio Ohtake

Visualization: Michio Murakami

Project administration: Fumio Ohtake

Funding acquisition: Michio Murakami and Fumio Ohtake

## Data availability

Data will be made available upon request.

## AI Statement

The authors used DeepL and ChatGPT to improve the English language of the manuscript. The manuscript was initially drafted by the authors, who critically reviewed and edited all AI-assisted revisions. The authors take full responsibility for the content of this publication.

