## Supplementary Materials for "Greater intergroup bias in vaccination attitudes among physicians than the general public"

Table S1. Unstandardized regression coefficients for intergroup bias, ingroup attitude, and outgroup attitude. Random effects are as follows: full sample—intergroup bias, between-person standard deviation ($\sigma_u$) = 2.45, residual standard deviation ($\sigma_e$) = 1.74, intraclass correlation coefficient (ICC) = 0.66; ingroup attitude, $\sigma_u$ = 1.85, $\sigma_e$ = 1.06, ICC = 0.75; outgroup attitude, $\sigma_u$ = 1.79, $\sigma_e$ = 1.11, ICC = 0.72; physician—intergroup bias, $\sigma_u$ = 2.48, $\sigma_e$ = 1.68, ICC = 0.68; ingroup attitude, $\sigma_u$ = 1.28, $\sigma_e$ = 0.91, ICC = 0.67; outgroup attitude, $\sigma_u$ = 1.83, $\sigma_e$ = 1.08, ICC = 0.74. Variance inflation factor: ≤ 1.55 for full sample; ≤ 2.59 for physicians.

| | Full sample | | | | | | Physician | | | | | |
|---|---|---|---|---|---|---|---|---|---|---|---|---|
| | Intergroup bias | | Ingroup attitude | | Outgroup attitude | | Intergroup bias | | Ingroup attitude | | Outgroup attitude | |
| | B (95% CI) | *P* | B (95% CI) | *P* | B (95% CI) | *P* | B (95% CI) | *P* | B (95% CI) | *P* | B (95% CI) | *P* |
| Agreement group (ref: disagreement) | 1.82 (1.72–1.91) | <0.001 | 1.49 (1.42–1.55) | <0.001 | −0.22 (−0.28–−0.16) | <0.001 | 3.44 (3.05–3.83) | <0.001 | 2.05 (1.85–2.26) | <0.001 | −1.39 (−1.64–−1.14) | <0.001 |
| Physician (ref: general public) | 1.66 (1.40–1.93) | <0.001 | 1.21 (1.02–1.41) | <0.001 | −0.49 (−0.68–−0.30) | <0.001 | - | - | - | - | - | - |
| Pediatrics (ref: internal medicine) | - | - | - | - | - | - | 0.99 (0.01–1.97) | 0.047 | 0.36 (−0.15–0.87) | 0.162 | −0.63 (−1.35–0.08) | 0.082 |
| Psychiatry (ref: internal medicine) | - | - | - | - | - | - | −0.23 (−1.07–0.62) | 0.599 | 0.05 (−0.39–0.49) | 0.819 | 0.28 (−0.34–0.89) | 0.375 |
| Surgery (ref: internal medicine) | - | - | - | - | - | - | −1.07 (−2.00–−0.14) | 0.025 | −0.45 (−0.93–0.04) | 0.072 | 0.63 (−0.06–1.31) | 0.072 |
| Other (ref: internal medicine) | - | - | - | - | - | - | −0.28 (−0.83–0.27) | 0.317 | −0.07 (−0.36–0.22) | 0.628 | 0.21 (−0.19–0.61) | 0.308 |
| Perceived benefits (distance) | 0.28 (0.25–0.32) | <0.001 | 0.12 (0.10–0.14) | <0.001 | −0.17 (−0.19–−0.15) | <0.001 | 0.14 (0.04–0.24) | 0.008 | 0.09 (0.03–0.14) | 0.002 | −0.05 (−0.12–0.01) | 0.109 |
| Perceived risks (distance) | 0.02 (−0.01–0.05) | 0.236 | −0.04 (−0.05–−0.02) | <0.001 | −0.07 (−0.09–−0.05) | <0.001 | 0.11 (0.01–0.20) | 0.025 | 0.04 (−0.01–0.09) | 0.104 | −0.07 (−0.13–−0.01) | 0.033 |
| Agreement (distance) | 0.60 (0.57–0.63) | <0.001 | 0.27 (0.25–0.29) | <0.001 | −0.31 (−0.33–−0.28) | <0.001 | 0.91 (0.81–1.00) | <0.001 | 0.42 (0.36–0.47) | <0.001 | −0.48 (−0.54–−0.41) | <0.001 |
| Age | 0.02 (0.01–0.02) | <0.001 | 0.03 (0.02–0.03) | <0.001 | 0.01 (0.00–0.01) | <0.001 | −0.01 (−0.03–0.02) | 0.592 | 0.00 (−0.01–0.01) | 0.946 | 0.01 (−0.01–0.02) | 0.431 |
| Man (ref: woman) | 0.04 (−0.11–0.19) | 0.589 | −0.14 (−0.25–−0.03) | 0.014 | −0.18 (−0.28–−0.07) | 0.001 | −0.91 (−1.75–−0.06) | 0.035 | −0.67 (−1.10–−0.23) | 0.003 | 0.24 (−0.37–0.86) | 0.434 |
| With Children (ref: without children) | 0.01 (−0.16–0.19) | 0.876 | 0.11 (−0.02–0.25) | 0.087 | 0.10 (−0.03–0.23) | 0.126 | −0.29 (−0.86–0.27) | 0.307 | 0.06 (−0.23–0.35) | 0.684 | 0.36 (−0.05–0.77) | 0.089 |
| No answer (ref: without children) | 0.17 (−0.67–1.00) | 0.697 | 0.12 (−0.50–0.73) | 0.708 | −0.04 (−0.64–0.57) | 0.908 | −0.73 (−2.45–0.99) | 0.402 | −0.40 (−1.30–0.49) | 0.374 | 0.33 (−0.92–1.58) | 0.604 |
| With partner (ref: without partner) | −0.20 (−0.38–−0.02) | 0.027 | −0.09 (−0.23–0.04) | 0.168 | 0.11 (−0.02–0.24) | 0.106 | 0.38 (−0.53–1.29) | 0.417 | 0.30 (−0.17–0.77) | 0.216 | −0.08 (−0.74–0.58) | 0.809 |
| No answer (ref: without partner) | −0.69 (−1.45–0.06) | 0.073 | −1.04 (−1.60–−0.48) | <0.001 | −0.35 (−0.89–0.20) | 0.212 | 0.32 (−1.71–2.36) | 0.755 | −0.05 (−1.11–1.01) | 0.923 | −0.38 (−1.86–1.11) | 0.618 |
| Influenza (ref: measles) | 0.04 (−0.04–0.11) | 0.315 | 0.13 (0.08–0.17) | <0.001 | 0.09 (0.05–0.14) | <0.001 | −0.23 (−0.47–0.01) | 0.061 | 0.02 (−0.11–0.15) | 0.797 | 0.25 (0.09–0.40) | 0.002 |
| HPV (ref: measles) | −0.01 (−0.08–0.07) | 0.808 | −0.06 (−0.11–−0.02) | 0.005 | −0.05 (−0.10–0.00) | 0.042 | −0.01 (−0.23–0.21) | 0.923 | 0.00 (−0.12–0.12) | 0.960 | 0.01 (−0.13–0.15) | 0.884 |
| COVID-19 (ref: measles) | 0.16 (0.09–0.24) | <0.001 | −0.03 (−0.08–0.01) | 0.151 | −0.18 (−0.23–−0.13) | <0.001 | −0.02 (−0.29–0.25) | 0.871 | 0.00 (−0.14–0.15) | 0.955 | 0.03 (−0.14–0.21) | 0.700 |
| Intercept | −2.33 (−2.64–−2.01) | <0.001 | 2.73 (2.51–2.96) | <0.001 | 4.95 (4.73–5.18) | <0.001 | −1.05 (−2.47–0.37) | 0.147 | 4.37 (3.62–5.11) | <0.001 | 5.38 (4.37–6.39) | <0.001 |

Table S2. Unstandardized regression coefficients for intergroup bias, stratified by vaccine type. Variance inflation factor: ≤ 1.57.

| | Influenza | | Measles | | HPV | | COVID-19 | |
|---|---|---|---|---|---|---|---|---|
| | B (95% CI) | *P* | B (95% CI) | *P* | B (95% CI) | *P* | B (95% CI) | *P* |
| Agreement group (ref: disagreement) | 2.79 (2.60–2.98) | <0.001 | 2.57 (2.36–2.77) | <0.001 | 2.54 (2.34–2.74) | <0.001 | 2.26 (2.07–2.46) | <0.001 |
| Physician (ref: general public) | 1.33 (1.03–1.63) | <0.001 | 1.55 (1.24–1.87) | <0.001 | 1.49 (1.18–1.81) | <0.001 | 1.08 (0.73–1.42) | <0.001 |
| Perceived benefits (distance) | 0.15 (0.09–0.22) | <0.001 | 0.25 (0.18–0.33) | <0.001 | 0.33 (0.26–0.40) | <0.001 | 0.23 (0.16–0.31) | <0.001 |
| Perceived risks (distance) | −0.02 (−0.08–0.04) | 0.541 | −0.07 (−0.13–−0.01) | 0.031 | −0.13 (−0.19–−0.07) | <0.001 | −0.08 (−0.15–−0.02) | 0.016 |
| Agreement (distance) | 0.82 (0.75–0.89) | <0.001 | 0.85 (0.78–0.93) | <0.001 | 0.71 (0.63–0.78) | <0.001 | 0.83 (0.76–0.90) | <0.001 |
| Age | 0.02 (0.01–0.02) | <0.001 | 0.02 (0.01–0.02) | <0.001 | 0.02 (0.01–0.02) | <0.001 | 0.02 (0.01–0.03) | <0.001 |
| Man (ref: woman) | 0.12 (−0.06–0.29) | 0.196 | 0.13 (−0.06–0.31) | 0.172 | 0.02 (−0.16–0.21) | 0.808 | 0.06 (−0.14–0.25) | 0.567 |
| With Children (ref: without children) | −0.07 (−0.28–0.14) | 0.517 | 0.06 (−0.15–0.27) | 0.584 | 0.06 (−0.17–0.28) | 0.629 | −0.12 (−0.35–0.11) | 0.311 |
| No answer (ref: without children) | −0.06 (−1.04–0.93) | 0.909 | 0.19 (−0.81–1.19) | 0.714 | 0.15 (−0.85–1.16) | 0.766 | 0.57 (−0.53–1.67) | 0.309 |
| With partner (ref: without partner) | −0.20 (−0.41–0.01) | 0.061 | −0.29 (−0.51–−0.07) | 0.009 | −0.21 (−0.44–0.02) | 0.071 | −0.10 (−0.34–0.13) | 0.377 |
| No answer (ref: without partner) | −0.54 (−1.41–0.33) | 0.228 | −0.74 (−1.67–0.18) | 0.114 | −0.40 (−1.34–0.54) | 0.409 | −0.92 (−1.88–0.03) | 0.059 |
| Intercept | −3.12 (−3.52–−2.72) | <0.001 | −3.38 (−3.82–−2.95) | <0.001 | −2.88 (−3.33–−2.44) | <0.001 | −2.88 (−3.32–−2.44) | <0.001 |

Table S3. Unstandardized regression coefficients for intergroup bias, ingroup attitude, and outgroup attitude (full sample), stratified by agreement and disagreement groups. Random effects are as follows: intergroup bias: agreement group, between-person standard deviation ($\sigma_u$) = 2.72, residual standard deviation ($\sigma_e$) = 1.14, intraclass correlation coefficient (ICC) = 0.85; disagreement group, $\sigma_u$ = 2.61, $\sigma_e$ = 1.33, ICC = 0.79; ingroup attitude: agreement group, $\sigma_u$ = 1.69, $\sigma_e$ = 0.73, ICC = 0.84; disagreement group, $\sigma_u$ = 2.05, $\sigma_e$ = 0.91, ICC = 0.83; outgroup attitude: agreement group, $\sigma_u$ = 1.94, $\sigma_e$ = 0.79, ICC = 0.86; disagreement group, $\sigma_u$ = 1.75, $\sigma_e$ = 0.98, ICC = 0.76. Variance inflation factor: ≤ 1.59 for agreement group; ≤ 1.92 for disagreement group.

| | Intergroup bias | | | | Ingroup attitude | | | | Outgroup attitude | | | |
|---|---|---|---|---|---|---|---|---|---|---|---|---|
| | Agreement group | | Disagreement group | | Agreement group | | Disagreement group | | Agreement group | | Disagreement group | |
| | B (95% CI) | *P* | B (95% CI) | *P* | B (95% CI) | *P* | B (95% CI) | *P* | B (95% CI) | *P* | B (95% CI) | *P* |
| Physician (ref: general public) | 1.71 (1.42–2.00) | <0.001 | −0.19 (−0.75–0.37) | 0.514 | 0.91 (0.73–1.09) | <0.001 | 0.71 (0.28–1.14) | 0.001 | −0.80 (−1.00–−0.59) | <0.001 | 0.88 (0.50–1.27) | <0.001 |
| Perceived benefits | 0.29 (0.26–0.31) | <0.001 | −0.30 (−0.34–−0.26) | <0.001 | 0.22 (0.20–0.23) | <0.001 | −0.04 (−0.07–−0.02) | 0.001 | −0.07 (−0.09–−0.06) | <0.001 | 0.26 (0.24–0.29) | <0.001 |
| Perceived risks | −0.11 (−0.13–−0.09) | <0.001 | 0.23 (0.20–0.26) | <0.001 | −0.01 (−0.02–0.01) | 0.389 | 0.20 (0.18–0.22) | <0.001 | 0.10 (0.08–0.12) | <0.001 | −0.01 (−0.03–0.01) | 0.326 |
| Agreement | 0.54 (0.51–0.57) | <0.001 | −0.26 (−0.31–−0.21) | <0.001 | 0.30 (0.28–0.32) | <0.001 | 0.01 (−0.03–0.04) | 0.651 | −0.24 (−0.27–−0.22) | <0.001 | 0.26 (0.23–0.30) | <0.001 |
| Age | 0.01 (0.01–0.02) | <0.001 | 0.01 (0.00–0.02) | 0.023 | 0.02 (0.02–0.02) | <0.001 | 0.03 (0.02–0.04) | <0.001 | 0.00 (0.00–0.01) | 0.056 | 0.02 (0.01–0.02) | <0.001 |
| Man (ref: woman) | 0.14 (−0.04–0.33) | 0.134 | −0.02 (−0.24–0.20) | 0.855 | 0.01 (−0.11–0.12) | 0.880 | −0.22 (−0.39–−0.05) | 0.012 | −0.13 (−0.26–0.00) | 0.048 | −0.20 (−0.35–−0.05) | 0.011 |
| With Children (ref: without children) | −0.01 (−0.22–0.21) | 0.947 | −0.03 (−0.29–0.24) | 0.850 | 0.09 (−0.04–0.23) | 0.186 | 0.15 (−0.05–0.36) | 0.140 | 0.10 (−0.06–0.25) | 0.210 | 0.18 (0.00–0.36) | 0.052 |
| No answer (ref: without children) | 0.13 (−0.92–1.17) | 0.811 | 0.39 (−0.81–1.60) | 0.522 | −0.27 (−0.92–0.38) | 0.413 | 0.30 (−0.63–1.23) | 0.527 | −0.40 (−1.14–0.34) | 0.290 | −0.06 (−0.87–0.76) | 0.894 |
| With partner (ref: without partner) | −0.25 (−0.48–−0.03) | 0.027 | −0.07 (−0.34–0.19) | 0.586 | −0.12 (−0.26–0.02) | 0.099 | −0.07 (−0.27–0.13) | 0.491 | 0.14 (−0.02–0.30) | 0.095 | 0.00 (−0.18–0.18) | 0.984 |
| No answer (ref: without partner) | −0.30 (−1.31–0.71) | 0.559 | −0.64 (−1.60–0.32) | 0.193 | −0.29 (−0.92–0.34) | 0.366 | −0.93 (−1.67–−0.18) | 0.015 | 0.01 (−0.71–0.73) | 0.977 | −0.30 (−0.95–0.36) | 0.376 |
| Influenza (ref: measles) | 0.10 (0.04–0.16) | <0.001 | −0.14 (−0.25–−0.03) | 0.016 | 0.20 (0.16–0.24) | <0.001 | 0.07 (−0.01–0.14) | 0.076 | 0.09 (0.05–0.13) | <0.001 | 0.21 (0.13–0.29) | <0.001 |
| HPV (ref: measles) | 0.04 (−0.02–0.10) | 0.228 | −0.03 (−0.14–0.09) | 0.652 | −0.01 (−0.05–0.03) | 0.681 | −0.01 (−0.09–0.07) | 0.753 | −0.05 (−0.09–0.00) | 0.032 | 0.02 (−0.06–0.10) | 0.647 |
| COVID-19 (ref: measles) | 0.17 (0.10–0.24) | <0.001 | 0.18 (0.06–0.29) | 0.003 | 0.04 (0.00–0.09) | 0.078 | 0.04 (−0.04–0.12) | 0.386 | −0.13 (−0.18–−0.08) | <0.001 | −0.14 (−0.23–−0.06) | <0.001 |
| Intercept | −3.72 (−4.15–−3.28) | <0.001 | 0.85 (0.40–1.30) | <0.001 | 1.85 (1.58–2.13) | <0.001 | 2.52 (2.17–2.86) | <0.001 | 5.61 (5.30–5.91) | <0.001 | 1.59 (1.29–1.90) | <0.001 |

Table S4. Unstandardized regression coefficients for intergroup bias, ingroup attitude, and outgroup attitude (excluding inattentive or ineligible responses). Random effects are as follows: intergroup bias, between-person standard deviation ($\sigma_u$) =2.51, residual standard deviation ($\sigma_e$) = 1.80, intraclass correlation coefficient (ICC) = 0.66; ingroup attitude, $\sigma_u$ = 1.73, $\sigma_e$ = 1.05, ICC = 0.73; outgroup attitude, $\sigma_u$ = 1.73, $\sigma_e$ = 1.11, ICC = 0.71. Variance inflation factor: ≤ 1.57.

| | Intergroup bias | | Ingroup attitude | | Outgroup attitude | |
|---|---|---|---|---|---|---|
| | B (95% CI) | *P* | B (95% CI) | *P* | B (95% CI) | *P* |
| Agreement group (ref: disagreement) | 1.98 (1.86–2.10) | <0.001 | 1.46 (1.38–1.53) | <0.001 | −0.45 (−0.53–−0.37) | <0.001 |
| Physician (ref: general public) | 1.66 (1.35–1.96) | <0.001 | 1.05 (0.84–1.26) | <0.001 | −0.63 (−0.84–−0.42) | <0.001 |
| Perceived benefits (distance) | 0.29 (0.26–0.33) | <0.001 | 0.13 (0.11–0.16) | <0.001 | −0.16 (−0.19–−0.14) | <0.001 |
| Perceived risks (distance) | 0.02 (−0.01–0.06) | 0.217 | −0.02 (−0.04–0.00) | 0.076 | −0.05 (−0.07–−0.02) | <0.001 |
| Agreement (distance) | 0.70 (0.66–0.74) | <0.001 | 0.34 (0.32–0.37) | <0.001 | −0.34 (−0.36–−0.31) | <0.001 |
| Age | 0.01 (0.01–0.02) | <0.001 | 0.02 (0.02–0.03) | <0.001 | 0.01 (0.00–0.01) | 0.004 |
| Man (ref: woman) | 0.15 (−0.04–0.34) | 0.125 | −0.08 (−0.21–0.05) | 0.227 | −0.23 (−0.35–−0.10) | <0.001 |
| With Children (ref: without children) | 0.03 (−0.19–0.26) | 0.769 | 0.11 (−0.04–0.26) | 0.163 | 0.08 (−0.08–0.23) | 0.333 |
| No answer (ref: without children) | 0.21 (−0.86–1.28) | 0.702 | −0.19 (−0.91–0.53) | 0.610 | −0.38 (−1.11–0.34) | 0.303 |
| With partner (ref: without partner) | −0.31 (−0.54–−0.08) | 0.008 | −0.13 (−0.29–0.02) | 0.093 | 0.18 (0.02–0.33) | 0.026 |
| No answer (ref: without partner) | −0.56 (−1.67–0.56) | 0.328 | −0.62 (−1.37–0.13) | 0.107 | −0.08 (−0.83–0.68) | 0.842 |
| Influenza (ref: measles) | 0.07 (−0.02–0.16) | 0.145 | 0.13 (0.08–0.19) | <0.001 | 0.07 (0.01–0.13) | 0.019 |
| HPV (ref: measles) | 0.02 (−0.07–0.12) | 0.654 | −0.05 (−0.11–0.00) | 0.065 | −0.07 (−0.13–−0.01) | 0.026 |
| COVID-19 (ref: measles) | 0.29 (0.19–0.39) | <0.001 | 0.00 (−0.06–0.06) | 0.927 | −0.26 (−0.33–−0.20) | <0.001 |
| Intercept | −2.47 (−2.88–−2.07) | <0.001 | 2.86 (2.59–3.13) | <0.001 | 5.22 (4.95–5.49) | <0.001 |
